# An mHealth-based social support program to improve antenatal care engagement and facility-based births in Uganda: A type I hybrid effectiveness-implementation clinical trial

**DOI:** 10.64898/2026.04.28.26351943

**Authors:** Esther C Atukunda, Godfrey R Mugyenyi, Jessica Haberer, Van T Nghiem, Elly B Atuhumuza, Peter Waiswa, Angella Musiimenta, Micheal Kanyesigye, Celestino Obua, Mark J Siedner, Lynn T. Matthews

## Abstract

**Background:** Ugandan women and their children suffer from high maternal and perinatal mortality, often due to low antenatal care (ANC) and skilled birth usage. We partnered with community members, women and the Ugandan Ministry of Health to formatively develop an intervention (*Support-Moms app*) to improve health education, engage social support networks, and augment access to ANC and delivery by a formal health care provider (HCP) for pregnant women in rural Uganda.

**Methods:** We conducted a type 1 hybrid effectiveness-implementation trial to test the effectiveness of the *Support-Moms* intervention. We enrolled 824 pregnant women (<20 weeks gestation) living in Southwestern Uganda and randomized them (1:1) to standard of care or *Support-Moms* intervention. The primary effectiveness outcome was completion of a HCP-led skilled birth (discharge card) and was analyzed as intention-to-treat. Secondary outcomes included number of ANC visits, institution-based delivery, social support, quality-of-life, mode of infant delivery, pre-term birth, birth weight, obstetric complications and deaths (maternal, fetal, newborn).

**Results:** A total of 1,216 women were screened, and 824 pregnant women enrolled (mean age ∼28 years; gestation at enrolment ∼13 weeks). Complete outcomes were available for 818 (99%). The *Support-Moms* intervention increased HCP-led skilled births compared to standard of care (93% vs 84%; OR 2.51, 95% CI 1.57–4.03, p<0.001). Women in the intervention group were more likely to achieve ≥4 ANC visits (84.1% vs 75.1%; OR 1.76, 95% CI 1.24–2.50, p=0.001) and less likely experience postpartum hemorrhage (9.1% vs 22.7%, OR 0.34, 95% CI 0.22–0.52, p<0.001) or for their neonates to require resuscitation (9.8% vs 13.7%, OR 0.69, 95% CI 0.45–0.99, p=0.001). Initiation of breastfeeding within an hour was higher (97.1% vs 71.7%, OR 1.76, 95% CI 1.15–3.44, p=0.001) and postnatal depression decreased (20.1% vs 27.1%, OR 0.68, 95% CI 0.49–0.94, p=0.019). More intervention participants reported adequate support, were birth-prepared and had a birth companion. There were no maternal deaths or differences in term births, birthweight, mode of delivery, perinatal mortality or other obstetric complications.

**Conclusions:** In rural Uganda, the Support-Moms mHealth-based, social-support intervention significantly increased HCP-led skilled births compared with routine care, while also improving ANC attendance, early breastfeeding initiation, birth preparedness, perceived social support and higher presence of companion at birth. Less women experienced PPH, neonatal resuscitation, and postnatal depression. Further evaluation should focus on the cost effectiveness and sustainability of this program.

**Trial registration:** Clinicaltrials.gov NCT05940831

## INTRODUCTION

Of the estimated 287,000 maternal deaths worldwide annually, 85% occur in low-and middle-income countries (LMICs) [1]. Antenatal care (ANC) and skilled births are mainstays of preventing maternal and perinatal morbidity and mortality [1–5]. Despite expanded availability of skilled birth attendants and referral health systems, Ugandan women still register low ANC attendance and skilled births, resulting in one of the highest maternal mortality ratios (MMR, 189/100,000) and perinatal mortality rates (41 deaths/1,000 births) in the world [6]. Lack of health education, social support, financial independence, decision-making autonomy regarding childbirth, birth preparedness, and perceived need for maternity services are predominant challenges to reducing pregnancy-related complications and MMR in these settings [7–11].

Mobile health (mHealth) interventions can promote healthcare utilization and ANC attendance by increasing knowledge, motivation, informing decision-making for women, and promoting partner involvement and access to social support [5, 12–16]. mHealth interventions that leverage social support can improve pregnancy experiences by decreasing anxiety and depression [17–20], increasing perinatal bonding [19], communication within social networks [20], and fostering coping mechanisms to help women overcome barriers to ANC attendance and skilled birth [20–23]. Whereas many mHealth interventions have been developed in Uganda [24–26], few have been in the reproductive health field [25, 27], and fewer have been evaluated at scale in the public sector [26, 28]. While the failure of impact has been attributed to a mismatch between the function, adaptability and need for mHealth interventions in some settings [5, 13], end-user designs that utilize iterative approaches in application development improve delivery success [29], and healthcare service utilization through internalize risks and benefits of health services [30].

To address these multi-dimensional gaps in care, we developed a user-centered mHealth-based, audio-SMS messaging application (*Support-Moms* app) aimed to share health-related information and engage social support networks to support pregnant women in rural Uganda to use available maternity services [31]. Using an iterative development approach, we conducted formative stakeholder interviews with women health care providers (HCPs) to identify preferred key ANC topics and characterize the preferred messaging intervention; developed content for SMS text messaging and audio messaging with the help of 4 medical experts based on the identified topics. The *Support-Moms* app (intervention), designed through partnership with an mHealth development company to engage social support networks, and pilot-tested in a randomized clinical trial was very promising [31, 32]. Nearly all intervention participants (98%, 39/40) had a skilled delivery compared to 78% (31/40) and 70% (28/40) in scheduled messaging and SOC groups, respectively.

To evaluate the program at scale, we performed a type 1 hybrid implementation-effectiveness trial [33, 34] comparing the *Support-Moms* intervention versus routine care in rural Uganda. We aimed to elucidate the benefit of this intervention to increase HCP-led skilled birth delivery, ANC uptake and maternal-fetal outcomes among rural, low literacy, and underserved populations in Southwestern Uganda.

## MATERIALS AND METHODS

### Ethics statement

This study was approved by the Institutional Review Committee of Mbarara University of Science and Technology and Uganda National Council of Science and Technology, and registered with clinicaltrials.gov (NCT05940831; https://clinicaltrials.gov/study/NCT05940831).

### Pilot data

In a randomized 3-arm pilot study (n=120) comparing standard of care (SOC), scheduled audio-SMS messaging, and scheduled messaging plus social supporter engagement facilitated via the *Support-Moms* app, we observed high intervention uptake, acceptability, and feasibility, with >80% women promptly receiving ≥85% of intended message [31, 32]. All women were able to identify at least 2 social supporters. All women whose social supporters engaged on the app (40/40) attended ≥4 ANC visits, compared to 83% (33/40) and 50% (20/40) of women receiving only scheduled messaging and SOC, respectively. Nearly all social support arm participants (98%, 39/40) had a skilled delivery compared to 78% (31/40) and 70% (28/40) in scheduled messaging and SOC groups, respectively. In qualitative interviews, women described the *Support-Moms* intervention as useful, actionable, and easy-to-use; it helped them learn, cope, prepare and take action within a friendly, trusted, and familiar environment. Involvement of partners and others helped them mobilize needed support. Our intervention is therefore hypothesized to improve awareness (through information transfer), and leverages existing CHWs, social networks and resources to encourage uptake, retention and use of available maternity care services within a community which largely depends on family and community networks to thrive [35]

### Study design and setting

We conducted a type 1 hybrid effectiveness-implementation trial to test the effectiveness of the *Support-Moms* intervention in two districts of southwestern Uganda. The sites were selected based on their geographic, socio-cultural, and institutional diversity, and high maternal mortality and morbidity. Both districts have publicly funded and operated facilities with active maternity care units. The local economy of these two districts is also largely based on subsistence agriculture, with both food and water insecurity being common [36, 37]; maternity services, including delivery, are largely provided free of charge through the public sector.

### Recruitment and enrolment of study participants

Women were eligible for inclusion if they: 1) were ≤ 20 (determined by last normal menstrual period-LNMP or ultrasound scan if available) and had not yet presented for ANC, 2) resided within 10km radius of all publicly funded maternity centers across Mbarara and Mitooma Districts, 3) were adults aged ≥ 18 years and/or emancipated minors (i.e. those <18 years of age who are pregnant), 4) reported access to a cell phone with reception in their home, 5) were able to identify at least two social supporters living within the study districts, and 6) were able to provide informed consent. Participants were recruited through community health workers (CHWs) in the study catchment areas.

CHWs notified study research assistants about potentially eligible participants, who then contacted and sought written informed consent before enrolment into the study. Participants were asked to identify two individuals from their existing social support network with whom they have had stable, long-term relationships and believe they would be available to help them during the pregnancy and study follow-up period. Eligible social supporters included spouses, relatives, CHWs and friends [23, 38] ≥18 years of age, who were aware that the study participant is pregnant, and owned a cell phone for personal use with self-reported reliable reception. Potential social supporters were excluded from the study if they were unable to use SMS or unwilling to receive SMS notifications. We emphasized selection of an existing partner that was aware of the pregnancy as one of the social supporters, alongside a friend, sibling, parent or CHW. Research assistants contacted social supporters from the intervention arm within two weeks of enrollment of pregnant women to confirm an active relationship at the time of their enrollment. Eligible social supporters were offered an explanation of study procedures and an opportunity to participate in informed consent. The study nurse informed consenting social supporters about the objectives of ANC and skilled delivery, as well as danger signs during pregnancy using standard MOH/WHO guidelines [3, 39]. All participants were allocated unique study numbers.

### Randomization

Prior to study initiation, the study statistician generated a randomization table, blinded to other study team members. Participants were stratified according to district and HC level, and randomly assigned to either intervention or control arms in a ratio of 1:1, in blocks of 10. Research assistants were informed of the arm assignment by the REDCap module after consent and at the time of enrolment. Study participants in the control group received Uganda’s MOH guidelines-based routine care and information giving [40]. The intervention group received the intervention described below.

### Intervention delivery and components

In our pilot study, we observed that knowledge gaps influenced women’s past and future decisions to not attend ANC and pursue unskilled home births [8, 41]. Women were also largely dependent on their significant others for economic provisions, which together with the existing gender and traditional norms, limited women’s ability and freedom to make family or health decisions to seek skilled care. We therefore developed a novel patient-centered mHealth-based, social support intervention using Andersen’s Healthcare Utilization Model to encourage and support women to use maternity care services in southwestern Uganda [8, 31, 32, 41, 42]. The final messaging prototype that included tailored SMS and audio health information was delivered by the *Support-Moms* application developed through a partnership with *iStreams-Uganda*, an application development company based in Mbarara that developed the app, and with an existing mHealth platform [43]. The unique multimedia design allowed women to be registered on the platform and be tracked throughout pregnancy and the post-partum period. Enrolled women received automated, scheduled SMS and audio messages, reminders, and notifications about upcoming appointments, as well as informational voice messages in their preferred language. The app includes a data collection platform [31], and stores information submitted in real-time directly from participant’s phone, thus allowing managers to access up-to-date data on process measures (e.g., automated messages sent and accessed) as well as intervention delivery and health outcomes. Fixed SMS data is stored in a secure cloud which is HIPAA compliant. *iStreams-Uganda* worked in partnership with *<u>Africas’ talking</u>*, a platform that facilitated access to a telco infrastructure that uses automated SMS, voice, airtime and other application programming interfaces (API)— mechanisms tested and successfully used during our pilot study. This automated technology for SMS [23, 44] and calls [29, 31] has also been used for other studies in Uganda.

Both SMS and audio messages were delivered at participants’ preferred time and day of the week to optimize intervention delivery. A weekly SMS reminder on the impending ANC appointment and expected date of delivery at their preferred time and day of the week, plus a day before the scheduled ANC visit was sent to study participants. Social supporters received weekly SMS notifications to motivate the pregnant women participant to present for scheduled ANC visits during the pregnancy as well as for delivery. Notifications to the two pre-identified social supporters provided information about the upcoming ANC visits and delivery due date during the study follow-up period. ANC appointment dates were automatically generated based on the provided last normal menstruation period (LNMP) and MOH guidelines [39] at enrolment. Social supporters were able to personalize the SMS content at enrollment, (default message will be “This is your reminder to assist your friend XXX attend her upcoming ANC visit due soon”). They were also advised to assist study participants with problems that may affect ANC attendance or facility delivery. The intervention was designed to build on existing supportive relationships of study participants within their communities. All women, and their social supporters received messages for at least 4 months, including the standard routine care provided at the community maternity centers.

### Data collection

Baseline participant characteristics were collected by study staff in both arms as well as among the social supporters in the intervention arm. We collected sociodemographic and clinical data on age, education, employment, socio-economic, marital status, religion, distance to the nearest health facility, history of home/facility birth gravidity, parity, gestational age, prenatal, antepartum high-risk morbidities, birth order, childbirth practices and relationships with HCPs. Baseline general and mental health was assessed using validated Hopkins Symptoms Checklist for depression, anxiety [45]. Postpartum depression was assessed using the validated Edinburgh postnatal depression scale [46]. We adopted the 6 items used in Uganda to assess personal and partner pregnancy desires, and 18 questions/statements reflecting 6 parenthood motives [47]. We assessed gender-based violence [48], and relationship power [49, 50] given its relationship with home births in Uganda [8]. We adopted and measured social support using a version of the Duke-UNC Functional Social Support Scale [51], a tool that has been widely used in Uganda [52]. We assessed the general health of women, including diagnosed NCDs, and measure Food insecurity using the Household Food Insecurity Access Scale (HFIAS) [53].

We collected outcome data in two ways: 1) through medical record review of the routinely provided ANC cards, postnatal discharge forms (where available), and records at the relevant health centres, and 2) through participant exit interviews 2-4 weeks following delivery, to enhance data completion, particularly for people who did not deliver at a facility. Skilled/facility births were collaborated with a facility record and or a postnatal discharge form. To additionally reduce risk of missing data, for participants who could not be contacted, we conducted home visits and interviewed next of kin. These survey data included the date and location of the birth, whether there was a skilled birth attendant present, mode of delivery (ie. vaginal vs C-section delivery), birth outcome; including preterm birth, maternal, fetal, newborn deaths, and any other complications of the birth (e.g obstructed labor, ruptured uterus, need for neonatal or maternal resuscitation, severe preeclampsia/ eclampsia, postpartum hemorrhage (PPH), maternal/newborn sepsis, and other infections), weight and height of newborn, number of ANC visits completed, use of breastfeeding and attendance at post-natal care. We also administered the Duke-UNC Functional Social Support Questionnaire [52] to measure reported social support received by women during pregnancy and childbirth.

### Statistical Analysis and Sample Size

Our primary outcome of interest was the proportion of women who had an HCP-led skilled birth delivery. We also explored additional secondary outcomes, including: a) number of ANC visits completed; 2) mode of infant delivery; 3) institution-based delivery; 4) presence of complications of birth; 5) child mortality; 6) maternal mortality; 7) preterm birth; 8) birth weight; 9) completion of postnatal care; 10) social support; 11) initiation of breastfeeding. Finally, we explored the role of social support as a moderating effect of the intervention through a pre-specified stratified analysis among women in the upper versus lower half of social support in the cohort, as measured by the Duke-UNC Functional Social Support Scale [51].

To test our primary effectiveness hypothesis, allowing for a two-sided type I error of 5%, 90% power, and assuming a 10% loss to follow up, we required 824 participants to detect a 10% difference in HCP-led skilled birth delivery between arms, assuming a 70% completion in the control arm, based on our pilot data and other similar studies in Uganda. [14, 15, 54]

We first summarized health-related and socio-demographic and clinical data between arms using standard descriptive statistics. We compared dichotomous outcomes between study groups by estimating crude odds ratios with 95% confidence intervals, and testing for differences between groups. We estimated *P*-values with chi-squared testing using a level of significance of 0.05. We compared continuous outcomes and estimated *P*-values using *t*-tests. All primary and secondary outcomes were analyzed using intention-to-treat analyses (although no participants were wrongly allocated a group). Although our study was fully randomized, the differences in baseline characteristics noted between study groups were assessed for confounding by fitting multivariable logistic regression models, and reported as per the revised CONSORT guidelines for reporting randomized trials [55]. We assessed for sub-group effects for the following characteristics by testing the significance of interaction terms in a multivariable regression model: 1) age (divided into ≤35 years and >35 years categories), 2) education (divided into primary and below, > primary), 3) decision-making autonomy (divided into not independent, independent), 4) parity (divided into 0, 1, 2, 3, 4 and ≥5), 5) inter-pregnancy interval of <2 years, 6) Food insecurity (divided into food secure, mildly food insecure, moderately food insecure, severely food insecure), 7) Regular income (No/Yes), 8) Household income (<150,000UGX, ≥150000UGX), 9) History of home birth (Yes/no), 10) intimate partner violence (Yes/No), 11) alcohol use (yes/no), 12) distance to health center (divided into within 2 km, 2–5 km, above 5 km), and 13) social support level (divided into low, medium, high). These sub-groups were not pre-specified but identified at data analysis stage when comparing different baseline characteristics across the two arms. A Mantel-Haenszel test was also done to control for each of these variables. All statistical analyses were performed using STATA version 17.0 (Statacorp, College Station, Texas, USA). An independent data safety monitoring board composed of members at the Mbarara University of Science and Technology, Makerere University and representatives from the study districts reviewed preliminary results at 50% (412) and 75% (618) of projected enrolment, as specified in the protocol, and recommended continuing study procedures.

### Funding

This publication was supported by the National Institutes of Health [NIH] National Institutes of Health; Eunice Kennedy Shriver National Institute of Child Health & Human Development (R01HD111692, awarded to ECA). The content is solely the responsibility of the authors and does not necessarily represent the official views of the NIH.

## RESULTS

### Participant characteristics

Between 1st May 2024 and 17th December 2025, a total of 1216 women were screened, of whom, 862 women were eligible, 824 were consented, enrolled and randomized in this trial (Figure 1). Of those excluded, 191 had initiated ANC visits, 153 had gestation age >20 weeks, 38 were not interested in the study. Out of the 824 enrolled, 818 completed study procedures, with a total of 6 (4 intervention, 2 control) participants lost-to-follow-up. Four hundred and eight participants received the intervention. The mean age was 28.1 (SD=6.3) years and 27.7 (SD=6.3) years) for the control and intervention arms respectively. Mean gestational age at enrolment (control 13.6 weeks; intervention 13.4 weeks), distance to facilities (control 4.2 km; intervention 4.3 km), education greater than primary (control 41.0%; intervention 41.7%), and HIV status (positive ∼9–10%) were similar across arms. Other demographic and clinical characteristics were similar between the two groups (Table 1).

**Figure 1.**
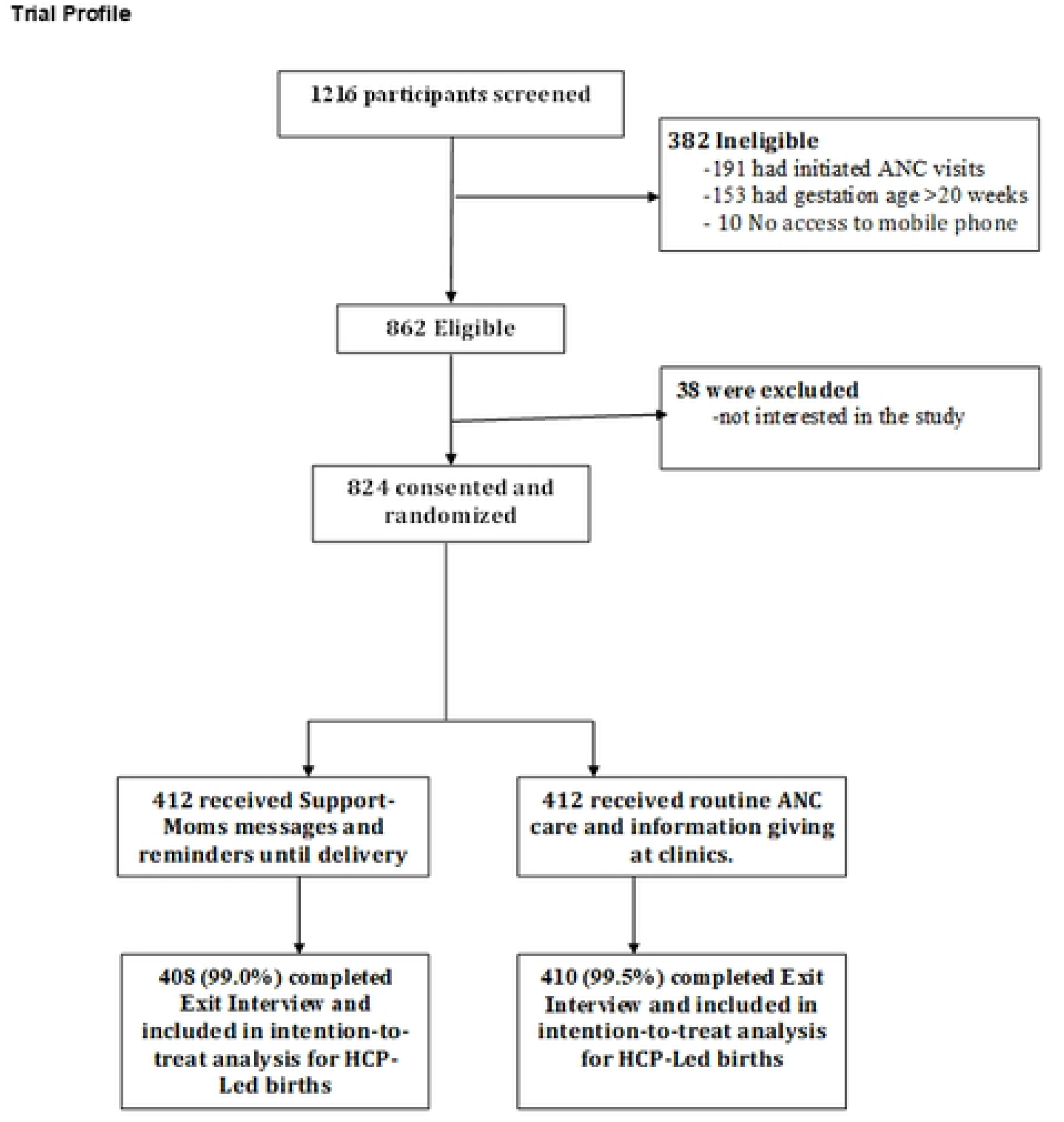

**Table 1:** Sociodemographic and clinical characteristics of participants by study arm.

| **Characteristics (Mean (SD) or n (%))** | **Standard care (n=412)** | **Support-Moms Intervention (n=412)** |
| --- | --- | --- |
| Age in completed years | 28.1 (6.3) | 27.7 (6.3) |
| Pregnancy partner age (years) | 33.4 (7.5) | 33.5 (8.2) |
| Educational attainment > primary | 169 (41.0) | 172 (41.7) |
| Mean gestation age at enrollment | 13.6 (4.0) | 13.4 (4.0) |
| Mean distance to publicly-funded facility (kms) | 4.2 (2.10) | 4.3 (2.24) |
| Distance to nearest health facility that offers ANC, EMOC/skilled delivery (kms) | 4.3 (2.10) | 4.3 (2.24) |
| Previous pregnancy interval< 1 year | 39 (9.5) | 25 (6.1) |
| Previous pregnancy interval< 2 year | 347 (84.2) | 362 (87.9) |
| Parity 0 | 56(13.6) | 69(16.7) |
| 1 | 134 (32.5) | 129 (31.3) |
| 2 | 78 (18.9) | 69 (16.7) |
| 3 | 70 (17.0) | 69 (16.7) |
| 4 | 44 (10.7) | 43 (10.4) |
| ≥5 | 30 (7.3) | 33 (8.0) |
| Household food insecurity<sup>a</sup> | 165 (40.0) | 165 (40.0) |
| Reported regular income | 46 (11.2) | 45 (10.9) |
| Household income >150,000 UGx/month | 149 (36.2) | 158 (38.3) |
| Median social support (IQR)<sup>b</sup> | 3(2.2-3.4) | 3(2.1-3.3) |
| Relationship of social supporter listed |  |  |
| Spouse | 309 (75.0) | 292 (70.9) |
| Mother | 11 (2.7) | 26 (6.3) |
| Friend | 41 (10) | 35 (8.5) |
| Sibling | 20 (4.9) | 20 (4.9) |
| CHWs | 1 (0.2) | 5 (1.2) |
| Other relative | 30 (7.3) | 34 (8.3) |
| Partner desired to have this pregnancy | 286 (69.4) | 273 (66.3) |
| Individual desired to have this pregnancy | 249 (60.4) | 232 (56.3) |
| Current pregnancy planned | 211 (51.2) | 201 (48.8) |
| Felt unhappy when found out about pregnancy | 142 (34.5) | 129 (31.3) |
| History of home birth<sup>c</sup> | 62 (18.2) | 58 (17.5) |
| History of perinatal death<sup>c</sup> | 43 (10.4) | 46 (11.2) |
| Mode of previous infant delivery<sup>c</sup> |  |  |
| SVD | 280 (82.4) | 262 (79.2) |
| C-Section | 60 (17.6) | 69 (20.3) |
| History of intimate partner violence | 135 (32.5) | 121 (29.4) |
| HIV serostatus |  |  |
| Positive | 37 (9.0) | 41 (10.0) |
| Negative | 370 (90.2) | 363 (89.0) |
| Unknown | 3 (0.7) | 4 (1.0) |
| Alcohol consumption in the last one year | 50 (12.2) | 52 (12.6) |
| Independent decision making | 112 (27.2) | 119 (28.9) |

<sup>a</sup> *HFIAS*>8 means severe food insecurity, <sup>b</sup> this score ranges from 1-4, with 4 indicating high levels of social support, <sup>c</sup> excludes prime gravid

The proportion of births attended by a healthcare provider (HCP-led skilled birth) was recorded in 379/408 (92.9%) in intervention vs 345/410 (84.1%) in control (OR 2.51 95% CI 1.57–4.03 p < 0.001; Table 2). The intervention also increased the proportion of women achieving ≥4 ANC visits, (OR 1.76 95% CI 1.24–2.50, p = 0.001). The intervention also reduced the need for resuscitation at birth [40/408 (9.8%) for intervention vs 56/410 (13.7%) in control; OR 0.69 95% CI 0.45–0.96 p = 0.037], and increased the likelihood of initiating breastfeeding within an hour after birth [intervention 396/408 (97.1%) vs control 294/410 (71.7%); OR 1.76 95% CI 1.15–3.44, p = 0.003]. Less women in the intervention (258/408, 63.2%) recorded one or more complications during labour/delivery compared to control (319/410, 77.8%; OR 0.51 95% CI 0.27–2.03, p = 0.027). Less postpartum hemorrhage (PPH) was reported for 37 out of 408 women in the intervention group (9.1%) versus 93 out of 410 (22.7%) for control (OR 0.34 95% CI 0.22–0.52, p < 0.001). Postnatal depression was also notably less among 82 women in the intervention group (20.1%) compared to 111 women (27.1%) in the control group (OR 0.68 95% CI 0.49–0.94, p = 0.019). Generally, adequate social support provided to women was significantly higher for 325 (79.7%) women in the intervention group compared to 289(70.5%) women in the control; OR 1.65 95% CI 1.19–2.29, p = 0.002. More women in the intervention group 142 (35.5%) versus 78 (19.0%) in the control group were adequately prepared for birth (OR 1.82 95% CI 1.29–2.46, p < 0.001), with a significant number of intervention women presenting at birth with a companion/social supporter (356 vs 337; OR 1.49 95% CI 1.01–2.20, p= 0.013). No statistically significant difference was seen in term births, birthweight, mode of delivery, perinatal deaths, miscarriage and maternal death. There was no maternal deaths in either arm.

**Table 2.**
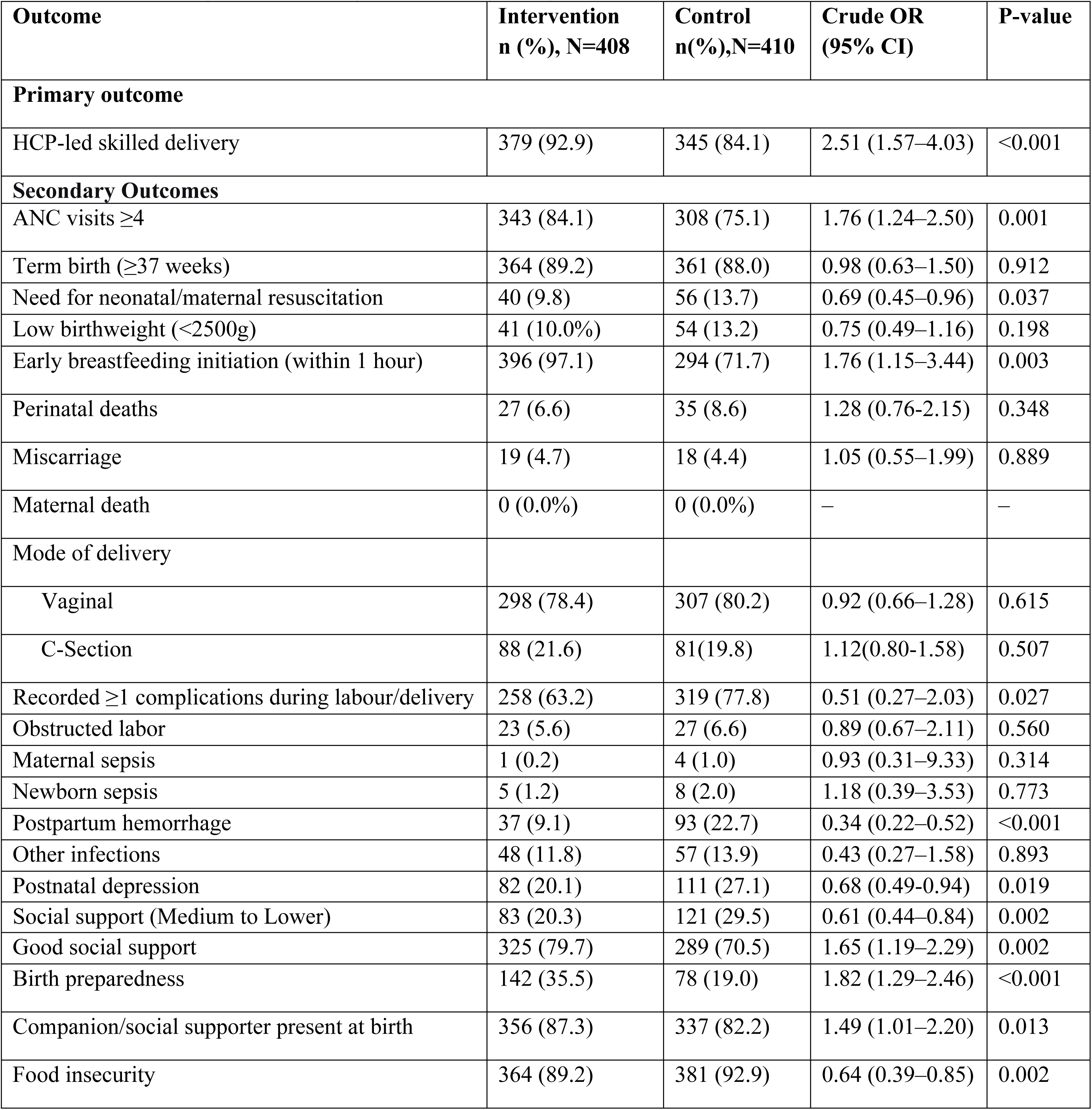
Primary and secondary outcomes by study arm.

In stratified analyses to assess for differences in the proportion of births attended by a healthcare provider (HCP-led skilled birth) within sub-groups, none of the sub-group-by-treatment interaction terms was significant except the inter-pregnancy interval <2 years (Figure 2). All of the point estimates showed higher proportions of HCP-led skilled birth in the intervention sub-groups compared to the control (Table 3). Thus, while the study was not powered to estimate effects within sub-groups, our results do not suggest differential effects of intervention within specific sub-groups of mothers.

**Figure 2.**
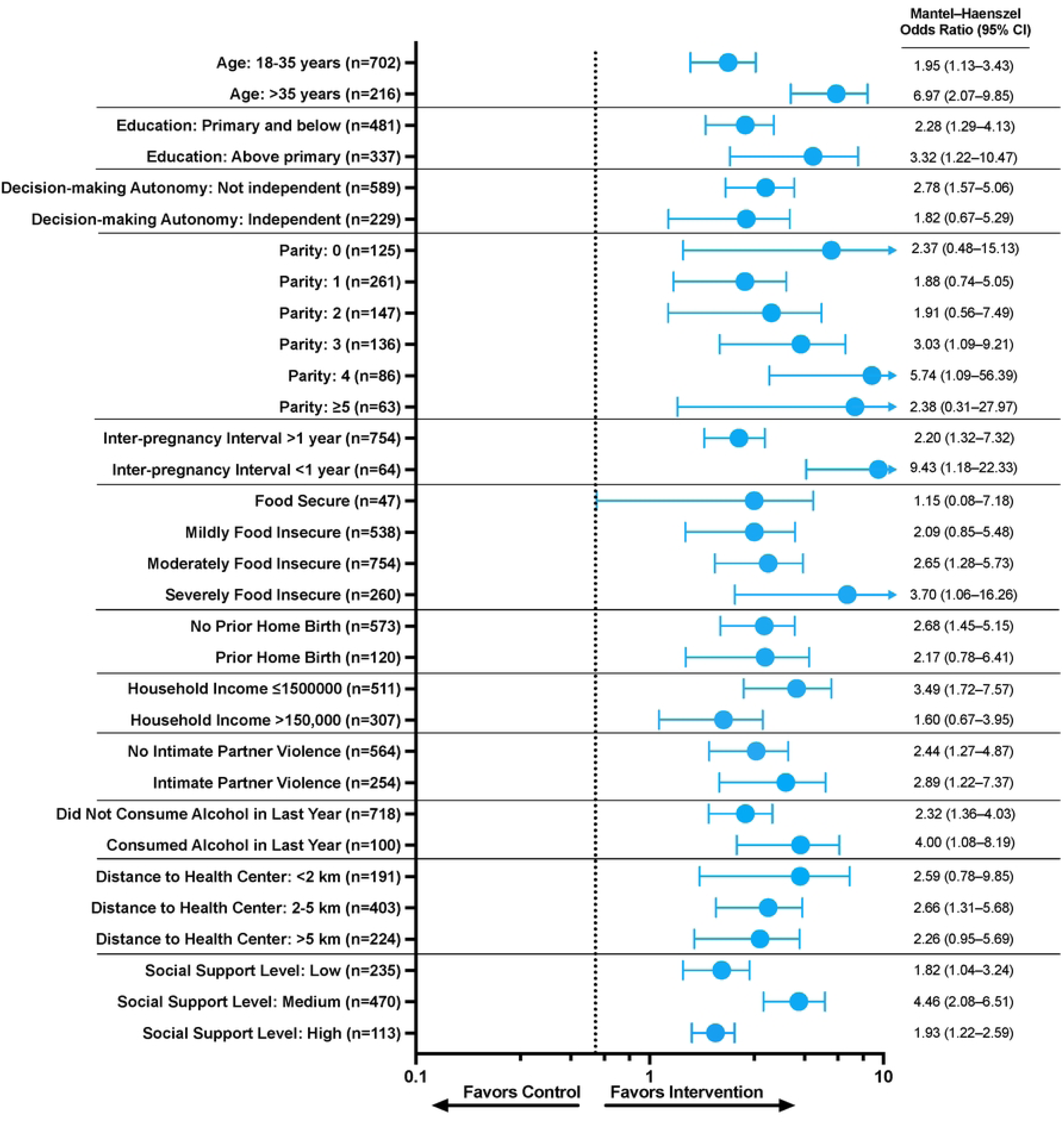

**Table 3.** Maternal baseline subcategories by study arms with skilled delivery.

| Characteristic | Intervention<br>n/N (%) | Control<br>n/N (%) | Mantel–Haenszel<br>odds ratio (95% CI) | p-value | p-value<br>(interaction) |
| --- | --- | --- | --- | --- | --- |
| **Age Category** |  |  |  |  | 0.057 |
| 18–35 years | 326/350(93.1) | 307/352 (87.2) | 1.95 (1.13–3.43) | 0.016* |  |
| >35 years | 55/58 (94.8) | 39/158 (67.2) | 6.97 (2.07–9.85) | 0.001* |  |
| **Education** |  |  |  |  | 0.500 |
| Primary and below | 218/238 (91.6) | 197/243 (81.1) | 2.28 (1.29–4.13) | 0.005* |  |
| Above primary | 163/170 (95.9) | 149/167 (89.2) | 3.32 (1.22–10.47) | 0.019* |  |
| **Decision-making autonomy** |  |  |  |  | 0.441 |
| Not independent | 270/290 (93.1) | 247/299 (82.6) | 2.78 (1.57–5.06) | <0.001 |  |
| Independent | 111/118 (94.1) | 99/111 (89.2) | 1.82 (0.67–5.29) | 0.244 |  |
| **Parity** |  |  |  |  | 0.869 |
| 0 | 66/69 (95.65) | 51/56 (91.07) | 2.37 (0.48–15.13) | 0.292 |  |
| 1 | 119/128 (92.97) | 115/133 (86.47) | 1.88 (0.74–5.05) | 0.186 |  |
| 2 | 65/69 (94.20) | 68/78 (87.18) | 1.91 (0.56–7.49) | 0.302 |  |
| 3 | 61/67 (91.04) | 51/69 (73.91) | 3.03 (1.09–9.21) | 0.033* |  |
| 4 | 39/42 (92.86) | 35/44 (79.55) | 5.74 (1.09–56.39) | 0.038* |  |
| ≥5 | 31/33 (93.94) | 26/30 (86.67) | 2.38 (0.31–27.97) | 0.406 |  |
| **Inter-pregnancy interval <6 mths** |  |  |  |  | 0.241 |
| No | 367/394 (93.15) | 335/393 (85.24) | 2.31 (1.40–3.88) | 0.001* |  |
| Yes | 14/14 (100) | 11/17 (64.71) | 9.10 (0.86–440.89) | 0.066 |  |
| **Inter-pregnancy interval <1 year** |  |  |  |  | 0.185 |
| No | 357/383 (93.2) | 318/371 (85.7) | 2.20 (1.32–3.72) | 0.003* |  |
| Yes | 24/25 (96.0) | 28/39 (71.8) | 9.43 (1.18–22.33) | 0.034* |  |
| **Inter-pregnancy interval <2 years** |  |  |  |  | 0.027 |
| No | 46/49 (93.9) | 42/65(64.6) | 8.40 (2.25–6.06) | 0.001* |  |
| Yes | 335/359 (93.3) | 304/345(88.1) | 1.80 (1.04–3.17) | 0.036* |  |
| **Food insecurity** |  |  |  |  | 0.756 |
| Food secure | 23/25 (92.0) | 20/22 (90.9) | 1.15 (0.08–7.18) | 0.917 |  |
| Mildly food insecure | 132/269 (49.1) | 137/269 (50.9) | 2.09 (0.85–5.48) | 0.108 |  |
| Moderately food insecure | 192/377 (50.9) | 185/377 (49.1) | 2.65 (1.28–5.73) | 0.009* |  |
| Severely food insecure | 62/130 (47.7) | 68/130 (52.3) | 3.70 (1.06–16.26) | 0.040* |  |
| **Regular income** |  |  |  |  | 0.399 |
| No | 339/364 (93.13) | 310/364 (85.16) | 2.31 (1.37–3.98) | 0.002* |  |
| Yes | 42/44 (95.5) | 36/46 (78.3) | 4.30 (1.01–25.48) | 0.048* |  |
| **Household income** |  |  |  |  | 0.335 |
| ≤1500000 | 236/250(94.4) | 213/261(81.6) | 3.49 (1.72–7.57) | <0.001 |  |
| Above 150,000 | 143/158 (90.5) | 132/149 (88.6) | 1.60 (0.67–3.95) | 0.291 |  |
| **Home birth history** |  |  |  |  | 0.915 |
| No | 264/281 (94.0) | 248/292 (84.9) | 2.68 (1.45–5.15) | 0.002* |  |
| Yes | 51/58 (87.9) | 47/62 (75.8) | 2.17 (0.78–6.41) | 0.039* |  |
| NA (First pregnancy) | 66/69 (95.7) | 51/56 (91.1) | 2.16 (0.40–14.45) | 0.368 |  |
| **Intimate partner violence** |  |  |  |  | 0.747 |
| No | 270/288 (93.8) | 238/276 (86.2) | 2.44 (1.27–4.87) | 0.008* |  |
| Yes | 111/120 (92.5) | 108/134 (80.6) | 2.89 (1.22–7.37) | 0.017* |  |
| **Ever consumed alcohol in last year** |  |  |  |  | 0.417 |
| Yes | 48/52 (92.3) | 36/48(75.0) | 4.00 (1.08–8.19) | 0.038* |  |
| No | 333/356 (93.5) | 310/362(85.6) | 2.32 (1.36–4.03) | 0.002* |  |
| **Distance to health center** |  |  |  |  | 0.955 |
| Within 2 km | 95/99 (96.0) | 81/92 (88.04) | 2.59 (0.78–9.85) | 0.121 |  |
| 2–5 km | 184/196 (93.9) | 174/207 (84.1) | 2.66 (1.31–5.68) | 0.007* |  |
| Above 5 km | 102/113 (90.3) | 91/111 (82.0) | 2.26 (0.95–5.69) | 0.066 |  |
| **Social support level** |  |  |  |  | 0.060 |
| Medium | 219/235 (93.2) | 198/235 (84.5) | 4.46 (2.08–6.51) | 0.001* |  |
| Lower | 111/119 (93.3) | 98/116 (84.5) | 1.82 (1.04–3.24) | 0.036* |  |
| High | 51/54(94.4) | 50/59(84.8) | 1.93 (1.22–2.59) | <0.001 |  |

## Discussion

In this randomized type 1 hybrid effectiveness-implementation trial conducted in rural Uganda, an mHealth-based, social-support intervention increased HCP-led skilled births compared with standard care. The intervention also led to higher ANC attendance (≥4 visits) and earlier initiation of breastfeeding, with concurrent reductions in postpartum haemorrhage and postpartum depression. There was improved social supporter engagement and support received by the women, birth preparedness, and higher presence of companion at birth. These findings suggest that programs that enhance engagement in maternal care and social support can meaningfully improve use of maternity services, birth preparedness, skilled births and some maternal-neonatal outcomes in similar settings.

Our patient-centred audio-SMS messaging intervention leveraged existing social support networks as cornerstones to deliver targeted information and stimulate instrumental social support needed to address practical and psychosocial barriers to facility-based delivery and ANC attendance prevalent in Uganda’s low-resource, rural settings [7–11, 54]. The observed effects on skilled birth attendance align with prior mHealth and hybrid interventions in low-resource settings that combine information, reminders, and social support to address barriers to care [26, 28]. Other studies have demonstrated that engagement of social support networks, partner involvement, and timely health information can reduce delays in seeking care and increase ANC attendance [56, 57], and institutional delivery [14, 15].

Regular audio-SMS messages can heighten awareness of danger signs, internalize risks and benefits of care seeking, reinforce birth preparedness, and strengthen informed decision-making to normalize facility-based birth [5, 12] [13, 58]. In our study, the magnitude of effect on skilled birth delivery appears similar to some previous trials in the region, which often report more modest gains when mHealth is implemented alone [24–26] [27]. The addition of a user-centred social-support component through active multi-modal mobile engagement may have amplified behavior change by tackling common informational, physical, social and economic barriers [7, 8, 54]. The scheduled phone-based messaging could have also increased consistent engagement, improved knowledge and awareness through targeted messages, increased motivation and social accountability via supporters [59],and cues to action [60], challenged societal negative beliefs [61] and strengthen birth preparedness. This pattern has been observed especially when interventions are well-directed and executed to provide accurate and relevant information [13, 62]. The effect seen on early breastfeeding initiation may reflect both improved postnatal guidance and greater caregiver involvement at birth. The reduction in postpartum haemorrhage and postpartum depression could reflect earlier, more frequent contact with social supporters, healthcare services and improved continuity of care, though residual confounding cannot be ruled out.

Another major component of the intervention was active engagement of social supporters within their existing networks (partners, family, friends, CHWs). Their role was meant to strengthen practical support, reduce transportation and cost barriers, and foster decision-making autonomy within households [51, 63, 64], key factors that have repeatedly been linked to higher uptake of healthcare services and infant care practices in sub-Saharan Africa [23, 38, 44, 65]. Prior mHealth interventions that specifically bolster social support have been documented to improve pregnancy experiences by decreasing anxiety and depression [17–20], increasing perinatal bonding [19] and communication within social networks [20]. These benefits are likely mediated through promoting existing beneficial family structure and social networks, which also in turn foster financial and emotional coping mechanisms to enable women to overcome socio-economic and physical barriers to care, such as food insecurity, transportation, and provision of delegated service to overcome competing priorities [20–23]. By leveraging existing networks and aligning with existing HCPs at nearby maternity care units, the intervention may have reduced fragmentation between community and facility-based care, facilitating smoother referral, timely care-seeking and better health outcomes [8, 23, 66–72]. Noteworthy, the rural, low-literacy, and resource-constrained setting in southwestern Uganda is characterized by strong reliance on family and community networks [35]. A low-cost intervention that foregrounds social support and user-centered content and approaches is plausibly better suited to this context to encourage uptake, end user engagement, retention and adoption than technology-only approaches [13, 62, 73],

This study had a number of strengths. We employed a randomized controlled design powered to detect a meaningful difference in a significant maternal health outcome. From our findings, we observed a 9% absolute increase in facility-based delivery with a formal health care worker (from 84.1% to 92.9%), with an absolute PPH difference of 13.6 percentage points [22.7% in control vs 9.1% in intervention; a relative reduction of approximately 60% (0.136 / 0.227)]. With Uganda’s current PPH rate at 25%, and approximate annual live births of 1.6–1.7 million [6, 8], our study suggests a reduction of about 60% of PPH with facility-based versus home-based birth, and that this intervention at national scale could translate into a reduction of about 144,000–155,000 home births and roughly 22,000–24,000 PPH cases per year in Uganda.

Although SMS alone is a convenient and lower cost approach to support healthcare interventions with higher delivery success, our intervention design focusing on phone access (versus ownership) within or outside the household. To enhance sustainability and scalability, we partnered with existing social networks to improve mobile phones access, and CHW to ease participant identification and recruitment. The provision of multiple messaging options accessible to any phone type such as voice and or text messages and active social networks involvement, may have been crucial to extend reach beyond the individual literate personal phone owners in sSA [74, 75]. Targeting Uganda’s low-literacy communities, and involvement of their available social networks may have therefore greatly benefitted women who have limited autonomy or financial independence to access timely care. To assess the robustness of the intervention to impact maternal and child health, we assessed multiple additional secondary outcomes, spanning uptake (ANC visits, facility delivery), maternal-fetal health outcomes (postpartum haemorrhage, postpartum depression, neonatal resuscitation, perinatal deaths, live births, birth weight, etc) to support process (social support, birth preparedness, birth companion, food security). Our type 1 hybrid design enabled simultaneous assessment of effectiveness and implementation, providing information for potential scale-up and full integration into routine care in similar settings. We used tracking procedures to assess outcomes and attained high retention and completion rates. Finally, to enhance generalizability, we conducted this study in multiple rural districts with diverse sociocultural contexts, similar health system tier system and network, enhancing external validity for similar settings.

This work also has a number of limitations. While the district selection aimed to capture diversity, generalizability may be limited to rural Uganda or similar settings where social networks play an important role in facilitating access to care [8]. Self-reported measures were used for some secondary outcomes that could introduce reporting bias. To mitigate this, we also conducted chart reviews of clinic data and used validated tools to measure postpartum depression, social support and food security. Involvement of the study at clinics could have contaminated results by improving care for all participants, although such an effect would be expected to bias our primary outcome comparison to the null. Notably, SMS were sent to intended recipients and women were routinely scheduled for ANC randomly and independently. The multiple secondary outcomes increase the risk of type I error, and multiple-testing adjustment was not conducted for these outcomes.

In summary, in a rural Ugandan setting, an mHealth-based, social-support intervention increased HCP-led skilled births compared with routine care, with concurrent improvements in ANC attendance, early breastfeed initiation, birth preparedness, and higher presence of companion at birth. There was also significant reductions in postpartum haemorrhage and postpartum depression. This multi-media mHealth strategy that couples informational reminders with social-support engagement mechanisms demonstrates great potential and promise in promoting maternity care utilization, warranting further evaluation of cost-effectiveness, other implementation outcomes, longer-term maternal-child outcomes and scalability for wider adaptation into routine maternal health programs within Uganda’s and similar settings. Considerations for scale-up may need to include data privacy, sustainability of mHealth messaging, and cost-effectiveness analyses, which are underway.

## Data Availability

The minimal data set is submitted alongside this manuscript

